# Topical Probiotics Decrease the Severity of Atopic Dermatitis. A Systematic Review and Meta-Analysis of Double-Blind, Randomized, Placebo Control Trials

**DOI:** 10.1101/2024.07.30.24311221

**Authors:** Elliot Flint, Nabeel Ahmad, Kevin Rowland, Charles Hildebolt, David Raskin

## Abstract

Atopic Dermatitis (AD) is a chronic skin disease that commonly appears during childhood but can present at any age. There are many reports showing that probiotics relieve AD symptoms in children. This systematic review and meta-analysis sought to determine the efficacy of topical probiotic treatment for AD in adult populations. A database search was conducted of peer-reviewed, double-blind clinical trials, and studies underwent a systematic exclusion and inclusion process, yielding four that met the criteria. Disease severity, as measured by a standardized scoring tool (SCORAD), was collected, and compared to placebo at two-week and four-week time points. All studies showed improvement in SCORAD in the treatment groups compared to baseline at all time points. Two showed significant decreases in SCORAD after two weeks of treatment, and three studies showed long-lasting improvement after four weeks of treatment. Interestingly, while each study showed a reduction in severity of AD at the two- and four-week time points, a pooled meta-analysis did not show a statistically significant difference between treatment and control at four weeks of treatment. Clinically, there may be benefits to topical probiotic usage as evidenced by the individual studies, more studies need to be performed including adults to show statistical significance.

## INTRODUCTION

Atopic dermatitis (AD) is an inflammatory skin disorder that often appears in early childhood, affecting approximately 8 - 12% of children^1^. AD can become a chronic condition, extending into adulthood, with 6 - 9% of U.S. adults affected^1^. An isolated cause has not been identified; however, a combination of family history, loss of function in the filaggrin (FLG) protein, which helps maintain the integrity of the skin, and exogenous environmental triggers, such as dust mites, heat, dry or humid climate, may play a role in its development^2^. The skin of AD lesions is characterized by microbial dysbiosis, with a reduction of diversity and overrepresentation of *Staphylococcus aureus*, correlating with increased lesion infection and flare-ups^3^. The normal microbiomes of the gut and the skin may produce molecules that inhibit the growth of *S. aureus*, and they interact with the immune system to downregulate inflammatory responses^4,5^. Given the relationship between the skin microbiome and AD, investigations into the use of adjuvant probiotics are warranted. Probiotics are living organisms that provide health benefits when consumed or applied to the body. Several systematic reviews have studied the effects of oral probiotics and their effect on decreasing AD severity in adult and pediatric populations. Such reviews explored various oral probiotic strains of Bifidobacterium, Lactobacillus, and/or Streptococcus with most trials concluding a reduction in the Scoring Atopic Dermatitis (SCORAD) index^6-12^.

There is a growing body of research investigating the effectiveness of probiotics on reducing the severity of AD. Most of these studies use oral probiotics and many test effectiveness only in children. There have been fewer studies using topical probiotics and testing their effectiveness in adults. Thus, a systematic review of randomized control trials of topical probiotics is needed to establish the validity of existing research regarding treating AD. To date, there has been no systematic review of topical probiotics for AD. In this systematic review, we explored the literature on topical probiotics’ role in decreasing AD in adult and pediatric populations. We performed a meta-analysis of the reported data to evaluate the effectiveness of the topical probiotics on reducing AD severity.

## MATERIALS AND METHODS

### Study Design

Web of Science and PubMed were searched using the terms (atopic dermatitis OR eczema) AND (probiotic* OR synbiotic*). After removing duplicates, the titles and abstracts of 979 articles were screened by two reviewers (EF and NA). Reviewers voted to exclude articles or advance them to the next stage of the review using the inclusion and exclusion criteria listed in Table 1. Reviewers’ decisions were blinded to one another and organized by the systematic review software Rayyan (Qatar Computing Research Institute). Any article that received that did not receive two votes to “exclude” was advanced to the next stage of the screening process (n=80). Article retrieval and screening of the full text of studies was done by four reviewers (EF, NA, DR, and KR). Each article was reviewed by two reviewers and voted to include or exclude based on the eligibility criteria. Conflicts between reviewer decisions were resolved by group consensus of the four reviewers. At the end of the screening process, only one article met the eligibility criteria for inclusion in the review. To expand the pool of eligible studies, the reviewers eliminated “Study subjects of 18 years or older” as part of the eligibility criteria. Articles were screened using the same process as above, and three additional articles were identified for inclusion in the review. Fig 1 highlights the preferred reporting items for systematic reviews and meta-analysis (PRISMA).

**Table I.** Inclusion and Exclusion Criteria.

| Inclusion | Exclusion |
| --- | --- |
| Subject age of 18 years or older<sup>1</sup> | Systematic Reviews/Review Articles |
| Randomized Control Trial | Published Before 2007 |
| Placebo | Receiving Other Treatments Targeted at Symptoms of Atopic Dermatitis |
| Use of SCORAD |  |
| Human Subjects |  |
| Topical Probiotic(s) |  |
| English Language |  |

<sup>1</sup>This criterion was removed for the second pass of the literature review

**Fig. 1.**
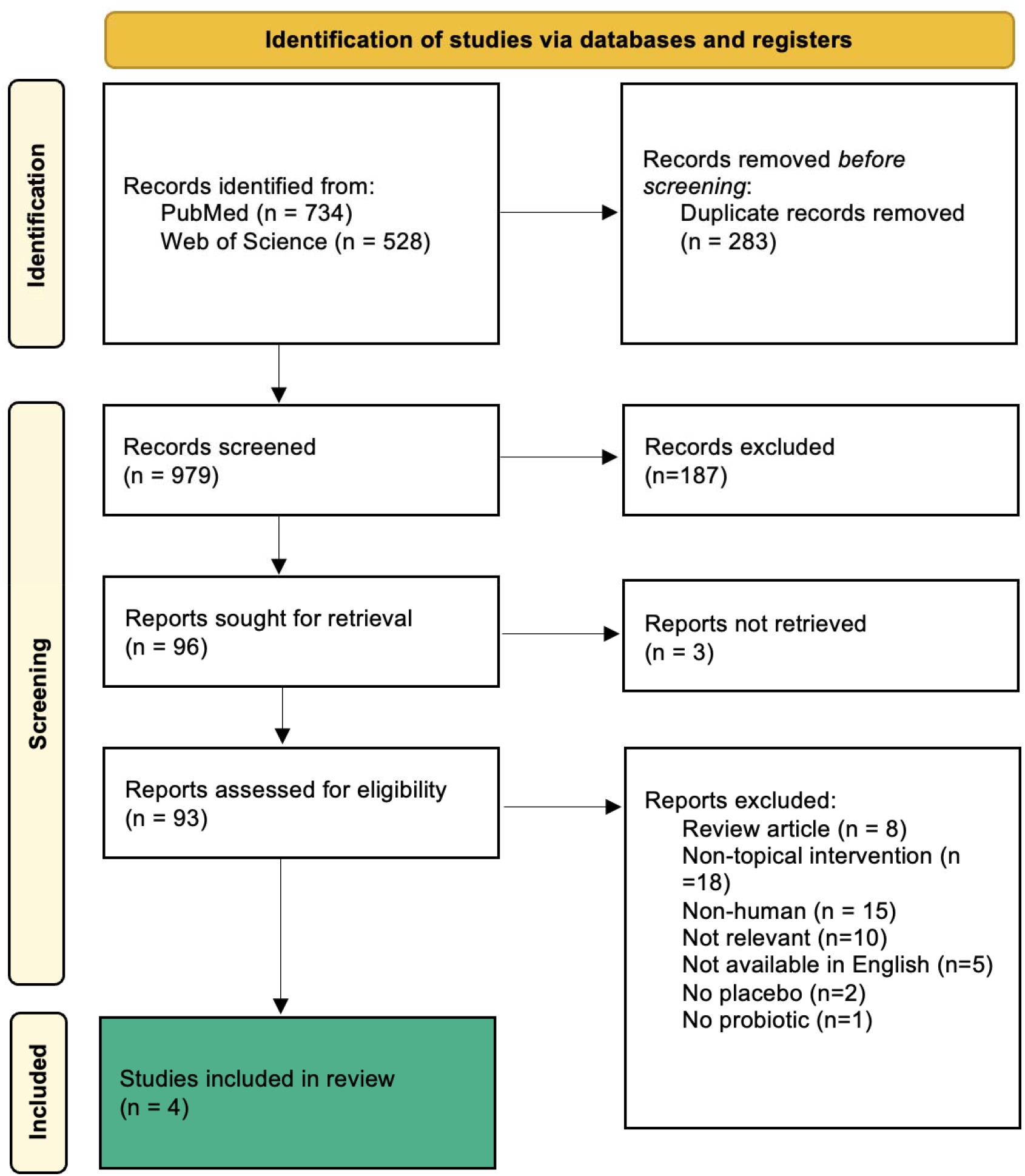
PRISMA flow diagram.

### Data Extraction

Data was extracted from the included articles by one reviewer (EF). Data extracted from each article included the probiotic(s) used, the study’s duration, the days at which treatment effects were measured, and the age of the study subjects. Raw data of SCORAD values were not included in the Axt-Gadermann study, so SCORAD values were obtained using the software NIH Image J to estimate values reliably based on graphs provided by the paper^13^.

### Statistical Analysis

Meta-analyses were performed for three time periods: (1) baseline, (2) two weeks, and four weeks. The intervals at which outcomes were measured varied slightly between studies. For our analysis, measurements taken on days 0 and 1 were grouped as “baseline.” Days 14 and 15 were grouped as “two weeks,” and days 28, 29, or 30 were grouped as “four weeks.” Four studies were used for the baseline assessments, two for the two-week assessments, and three for the four-week assessments. The standardized difference in means was used as the effect size index. Because we assumed that the studies in each analysis represented a random sample from the universe of potential studies, we employed a random-effects model for each analysis, with each analysis being used to make an inference to the universe of potential studies. In support of our using a random-effects model, we provide the following two statements from a book on meta-analysis. “When studies are pulled from the literature, a random-effects model should be used because common sense indicates that the true effect size varies across studies”^14^; and “This model assumes that the studies in the analysis are representative of a universe of comparable studies, and that the results of the analysis will be generalized to that universe^14^.” In addition, one of our goals was to create a prediction interval for each time period, and the creation of a prediction interval requires the use of a random-effects model. As indicated in another recently published book^15^, a prediction interval includes the true effect size for 95% of all populations in the universe.” In this book, the importance of prediction intervals in meta-analyses is stressed,^15^ and an article^16^ is cited that makes a plea for routinely presenting prediction intervals in meta-analyses; therefore, to help interpret the results of our meta-analysis, a prediction interval (which is based upon the random-effects model) was calculated for each time. “The prediction interval reflects the variation in treatment effects over different settings, including what effect is to be expected in future patients…”^16^ “(More accurately, in 95% of all meta-analyses the mean effect size will fall within the confidence interval.)”^14^ As explained in this cited book: The 95% confidence interval for a fixed-effect model tells us that the mean prevalence in the set of studies falls with this range, and the 95% confidence interval for a random-effects model tells us that the mean in the universe of comparable populations falls with this range; whereas, the 95% prediction interval tells us that the prevalence in any single population can be as low or as high as the lower limit and the upper limit of the interval^14^.

For our meta-analysis, the standardized mean difference (SMD) for a fixed-effect model was calculated with Hedges g statistic,^17^ and the heterogeneity statistic was used to calculate the summary standardized mean difference for a random effects model^18^. “If the value 0 is not within the 95% confidence interval (CI), the SMD is statistically significant at the 5% level (P < 0.05)^19^.” “Cohen’s rule of thumb for interpretation of the SMD statistic is: a value of 0.2 indicates a small effect, a value of 0.5 indicates a medium effect and a value of 0.8 or larger indicates a large effect^19^.”

Alpha was set at 0.05; however, Amrhein et al. emphasized that the clinical importance of findings, not their statistical significance, be emphasized,^20^ and for the interpretation of results of our meta-analysis, the clinical importance of findings are emphasized. For our meta-analysis, the standardized mean difference (SMD) for a fixed-effect model was calculated with Hedges g statistic,^17^ and the heterogeneity statistic was used to calculate the summary standardized mean difference for a random effects model^18^. “If the value 0 is not within the 95% confidence interval (CI), the SMD is statistically significant at the 5% level (P < 0.05)^20^.” “Cohen’s rule of thumb for interpretation of the SMD statistic is a value of 0.2 indicates a small effect, a value of 0.5 indicates a medium effect and a value of 0.8 or larger indicates a large effect^19^.” We performed our meta-analysis with Comprehensive Meta-Analysis Version 4^21^.

## RESULTS

Our search strategy yielded four articles that met the inclusion criteria^22-25^ (Table 2). Articles tested the use of probiotics in treating atopic dermatitis, although different timelines were used for testing.

**Table II.** Summary of all included articles.

| Study | Probiotic | Duration of Study/<br>Days<br>SCORAD<br>Measured | n / Age Range<br>of Subjects in<br>years<br>(placebo,<br>treatment) | Percent Change from<br>Baseline in SCORAD | Additional Findings |
| --- | --- | --- | --- | --- | --- |
| Noll et al, 2021 | Mixture<br>(*Bifidobacterium breve*,<br>*Bifidobacterium animalis lactis*,<br>*Lactobacillus casei*,<br>*L. gasseri*,<br>*L. planatarum*,<br>*L. rhamnosus*) | 2 weeks/<br>0,9,11,14 | 7,7 / 5+ | Synbiotic baths:<br>-40.7%, -49.7% and -56.8%<br>on days 9, 11, and 14,<br>respectively.<br>Placebo: -2.3%, +1.3%, and<br>+7.1% on days 9, 11, and 14,<br>respectively. | Analysis of skin flora showed successful colonization with<br>bacteria from the probiotic treatment, but the abundance of<br>*Staphylococcus aureus* did not significantly change.<br>Subjects in probiotic group subjectively reported less pruritis and<br>less dryness at the end of study compared to control group. |
| Butler et al, 2020 | *Lactobacillus reuteri* DSM17938 | 8 weeks/<br>0,28,56 | 17,17 /<br>19-66, 19-57 | Probiotic ointment:<br>-27.7% and -45.7% on days<br>28 and 56, respectively.<br>Placebo: -31.9% and<br>-41.0% on days 28 and 56,<br>respectively. | Improvement of skin dryness in the probiotic group was the most<br>pronounced change during the study. |
| Gueniche et al, 2008 | *Vitreoscilla filiformis* | 4 weeks/<br>0,15,29 | 25,26 /<br>6-70, 6-70 | Probiotic cream:<br>-29.7% and -56.1% on days<br>15 and 29, respectively.<br>Placebo: -7.8% and -16.3%<br>on days 15 and 29,<br>respectively. | Probiotic group showed significantly greater reduction in<br>SCORAD when compared to control at days 15 and 29.<br>Skin barrier function <u>was measured</u> using the biomarker TEWL.<br>There was a significant reduction in TEWL in both groups from<br>their baseline values at the end of the study, but there was no<br>significant difference between the probiotic vs control.<br>The group treated with probiotic had decreased colonization of<br>*Staphylococcus aureus* and *Escherichia coli* compared to control<br>at the end of the study, but the finding was not statistically<br>significant. |
| Seite et al, 2017 | *Vitreoscilla filiformis* (LRP-VFB) | 4 weeks/ 1,28 | 22,24 / 0-63, 0-63 | Probiotic cream:<br>-15.5% on day 28.<br>Placebo: +32.0% on day 28. | The probiotic group had significantly increased levels of<br>*Xanthomonas* at the end of the study compared to control.<br>Colonization with *Staphylococcus aureus* increased in the control<br>group, but not the treatment group. The difference was not<br>statistically significant. |

### SCORAD

SCORAD is a standardized scoring tool to rate the severity of atopic dermatitis by physician and patient ratings of pruritis, dryness, and redness. Higher SCORAD values indicate more severe symptoms, whereas a low SCORAD indicates milder disease severity. In the articles that were included in our analysis, the percent change in SCORAD demonstrated reduced eczema severity with the use of probiotics in all four studies (Fig 2a), whereas placebos only reduced eczema severity in two of the four studies (Fig 2b). The percent change in SCORAD from the probiotic groups was subtracted from the percent change in placebo at each respective time point and graphed; positive values indicate that the probiotics outperformed the placebo at that charted point (Fig 2c). This figure demonstrates how probiotics outperformed placebos across all studies, except for the Butler et al. study at 28 days. It should be noted, however, that the Butler study showed a benefit from the probiotics compared to placebo later in the study at 56 days.

**Fig. 2.**
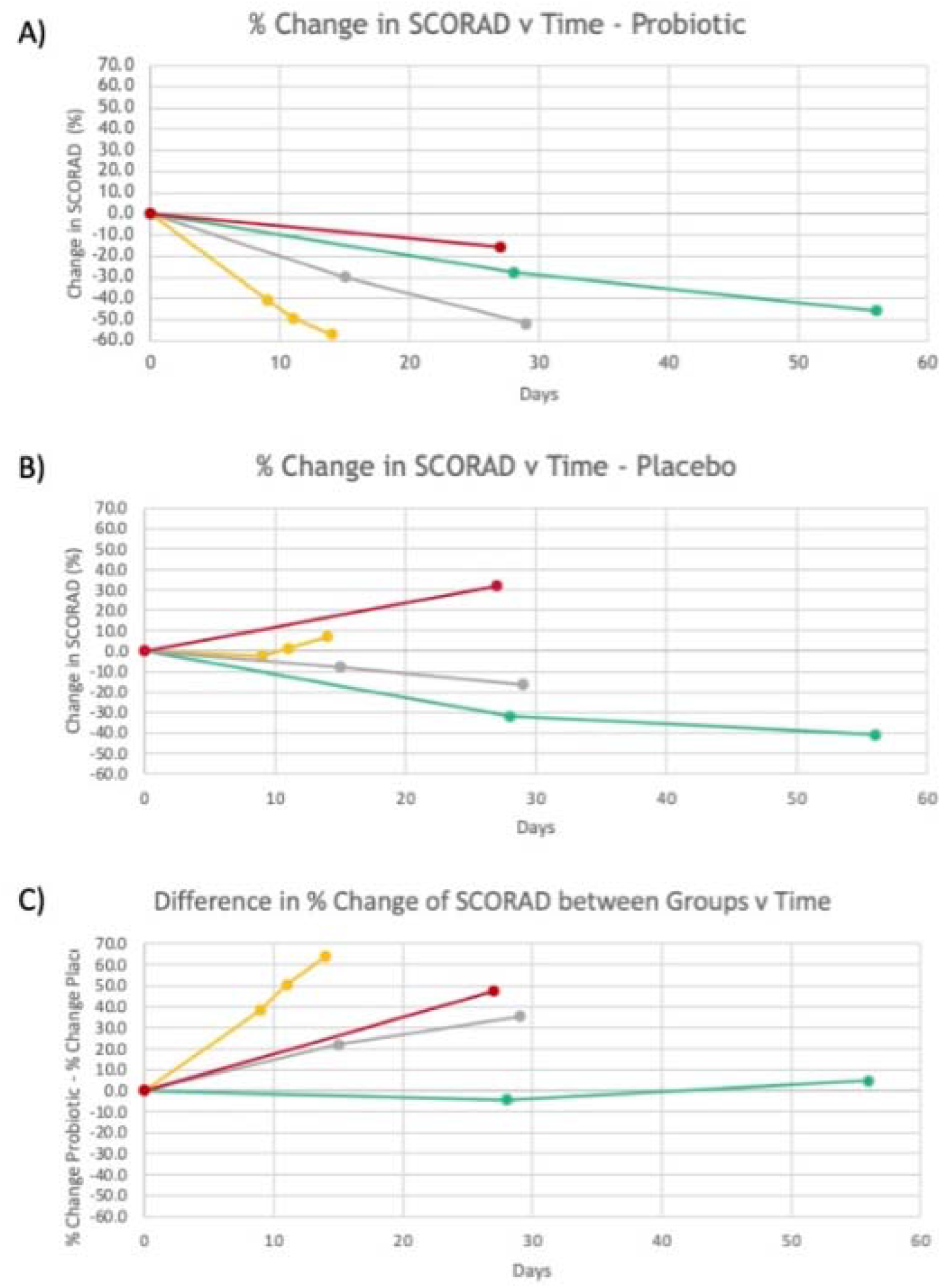
Percent Change from baseline in SCORAD vs time. A) Percent change in SCORAD vs time for probiotics for all studies. B) Percent change in SCORAD vs time for placebo for all studies. C) Difference in percent change in SCORAD between groups vs time for all studies.

#### Meta-analysis

Meta-analyses were performed for the placebo and probiotic treatment groups. For the baseline data, the mean effect size is 0.372 with a 95% confidence interval of -0.021 to 0.765 (Fig 3a). The mean effect size in the universe of comparable studies could fall anywhere in this interval. The Z-value tests the null hypothesis that the mean effect size is zero. The Z-value is 1.854 with p = 0.064. Using a criterion alpha of 0.050, we cannot reject this null hypothesis of no effect. If the true effects are normally distributed (in g units), an estimate of the prediction interval is -0.889 to 1.633 (Fig 3a). The true effect size in 95% of all comparable populations falls in this interval.

**Fig. 3.**
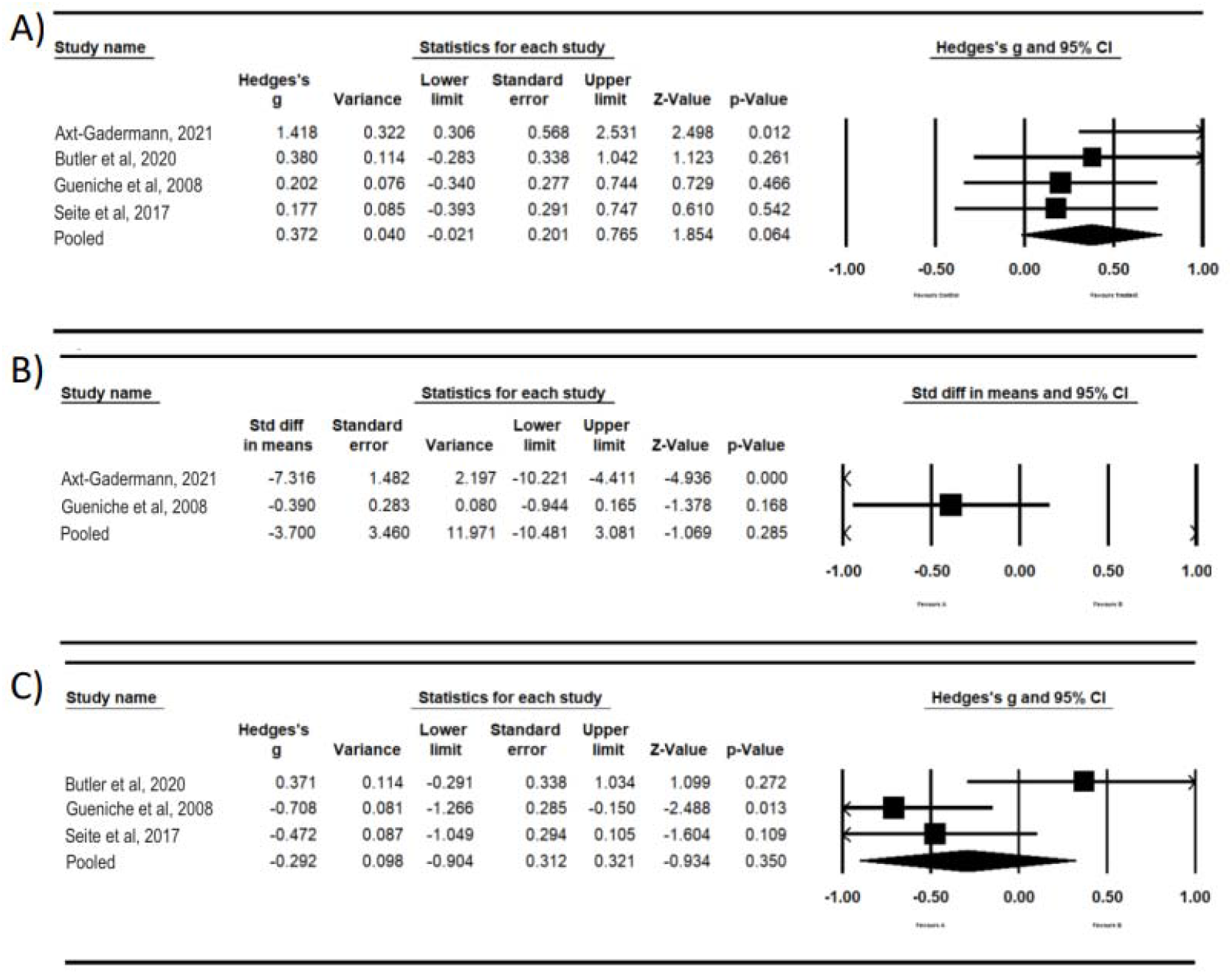
Summary values for dermatitis, (A) Baseline values for four studies and the pooled values for the random effects model. (B) Two-week values for two studies and the pooled values for the random effects model. (C) Four-week values for four studies and the pooled values for the random effects model.

These calculations for baseline data included only four studies. It has been suggested that ten studies are a useful minimum for a meta-analysis15. Therefore, estimates of heterogeneity based on less than ten studies may not be reliable. A prediction interval and other functions could not be performed for the two-week data because a minimum sample size of 3 is required, and for the two-week assessments, there were only two studies. For the two-week data, the mean effect size is -3.700 with a 95% confidence interval of -10.481 to 3.0815 (Fig 3b). The mean effect size in the universe of comparable studies could fall anywhere in this interval. The Z-value is -1.069 with p = 0.285, which does not reject the null hypothesis of no effect. For the four-week data, the mean effect size for the three studies is -0.292 with a 95% confidence interval of -0.904 to 0.321 (Fig 3c). The mean effect size in the universe of comparable studies could fall anywhere in this interval. The Z-value is -0.934 with p = 0.350. Using a criterion alpha of 0.050, we cannot reject this null hypothesis of no effect. An estimate of the prediction interval is -7.221 to 6.637 (Fig 4b). The true effect size in 95% of all comparable populations falls in this interval. These calculations for four-week data included only three studies; therefore, heterogeneity estimates may not be reliable.

**Fig. 4.**
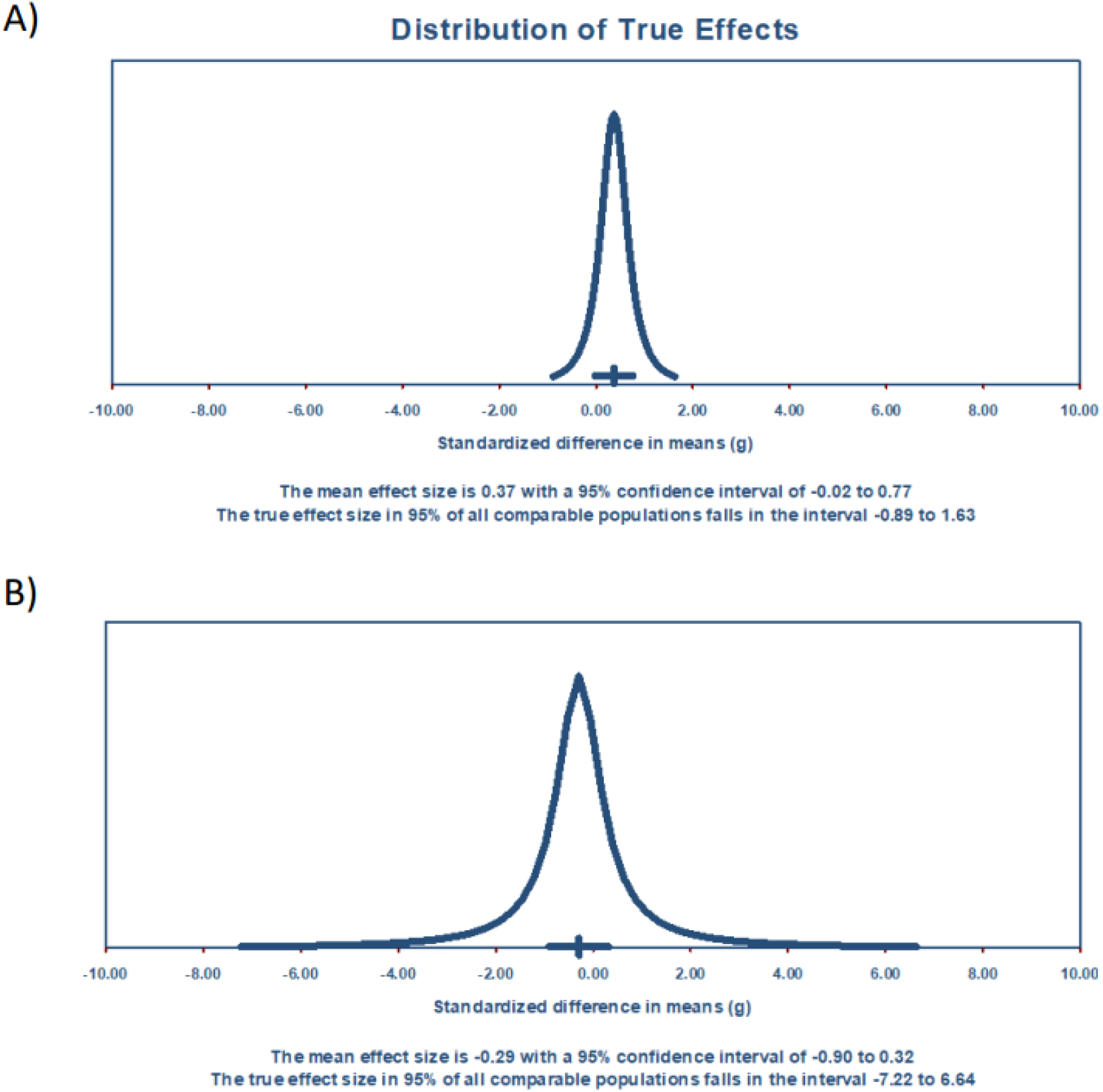
Prediction intervals for dermatitis at baseline and at four weeks—a two-week prediction model could not be created because a minimum sample size of 3 is required, (A) Prediction interval for baseline values. (B) Prediction interval for four-week values.

To summarize the results of our assessments, none of our meta-analyses rejected the null hypothesis of no effect (p ≥ 0.064). The prediction interval at baseline indicated that with treatment, many future patients would experience benefits; however, at four weeks, the prediction interval indicated that it would be essentially a coin toss as to whether a future patient would experience a benefit. A limitation of our assessments is that our sample sizes for baseline, two weeks, and four weeks were small—respectively, four studies, two studies, and three studies.

## DISCUSSION

AD lesions show an increased abundance of S. aureus and reduced diversity of the skin microbiome3,5. Due to a lack of long-term, effective AD treatments, there has been interest in modifying the skin microbiome using probiotics. Probiotics are easily assembled, relatively inexpensive, and can be purchased without a prescription, representing a practical avenue of treatment for individuals afflicted with AD. We performed a systematic review and meta-analysis of studies that investigated the use of topically applied probiotics for AD treatment. Overall, only a few studies met our inclusion criteria. The included studies demonstrate a benefit of probiotic application in treating atopic dermatitis as measured by a standard scoring system (SCORAD); however, that benefit varied at the different time points measured.

This systematic review had multiple challenges. Initially, we tried to perform a review of AD in adults, but there were not enough studies to carry that out. We modified the criteria to accept studies that included both children and adults. Even so, only four studies met the criteria. These studies used different probiotic formulations and different study periods. The meta-analysis showed a beneficial effect at two weeks but not at four weeks. The one study with enough participants to show the non-effect at four weeks showed a beneficial effect at eight weeks, suggesting that topical probiotics are effective, but effects may be better appreciated in studies of longer durations. Until more clinical trials are performed using standardized treatments and treatment lengths, it will be challenging to determine the effectiveness of topical probiotics. The one study that showed little effect on SCORAD for up to four weeks used a probiotic with only a single organism, *Lactobacillus reuteri*^23^. Two of the other studies used *Vitreoscilla filiformis*, and one study used a symbiotic, a mix of several bacterial species, along with a prebiotic (Table 1). Future studies should focus on probiotics that show effectiveness, and while there are only two studies, *V. filiformis* is promising as a probiotic treatment for AD^24,25^.

Atopic dermatitis commonly affects infants and young children, but adults are affected as well, with as many as 10% of adults reporting eczema^26,27^. The gut microbiome has been linked with skin health and may be important in preventing inflammatory processes associated with AD^28,29^. Probiotics may interfere *with S. aureus* growth through directly killing *S. aureus*, or occupying niches in the skin preventing *S. aureus* from colonizing. Decreasing *S. aureus* growth on the skin could increase diversity and reduce inflammation. Clinical trials using oral probiotics to modify the gut microbiome have been performed, but in adults, oral probiotics have shown mixed effects^30,31^. Using oral probiotics on those with mature gut microbiomes might not be as effective compared to using them for infants or young children. Our systematic review evaluated topical probiotics to determine whether there was an effective treatment for adults.

A recent systematic review analyzed many AD treatments, including oral and topical probiotics, topical emollients, biologics, pharmaceuticals, and many other therapies^30^. They found that treatments across the spectrum had positive effects on AD outcomes. Oral probiotics generally improved AD severity in children and adults, although some studies in children showed no effect. They found that topical probiotics generally improved AD, but they also included non-clinical trials in their results. They also found that there were significant benefits from biologics. Our systematic review and meta-analysis used only double-blind, randomized clinical trials, and all studies we analyzed included adults. We found that while there was a general reduction in SCORAD from topical probiotic use in three out of four studies, we could not reject the null hypothesis, that the treatment had no effect at four weeks. This is consistent with other research. Greenzaid et al. found that most of the reported studies show a positive effect on SCORAD or other indicators of AD, but there were also studies showing no effect. Many treatment studies show a benefit to AD patients, but the mixed results show that there is a need to determine which treatments work, why they work, and the appropriate length of treatment. For probiotics, there are a variety of different bacteria that are administered, and there are a variety of vehicles to carry the bacteria. Future studies need to determine the most effective treatments.

An advantage of topical probiotic treatment is that it is low-cost, easy to administer, and has few adverse effects. Using biologics or pharmaceuticals will be more costly. Oral administration of probiotics carries a greater risk of non-adherence to treatment. All four clinical trials we identified that used topical probiotics in randomized, double-blind studies show positive effects over the length of the trial. Our meta-analysis showed that this treatment is only effective over the first two weeks of treatment. Because endpoints varied between studies, the longest common endpoint, four weeks, was analyzed and showed no significant effect. We recommend that additional clinical trials be performed using the treatments that look successful (break out which probiotics were successful vs not), using more patients over a longer period to determine treatments that will be effective enough for widespread use.

## Data Availability

All data produced in the present work are contained in the manuscript

## IRB approval status

n/a

